# The Sleep-Wake Classification Performance of Pediatric-Trained Machine Learning Algorithms for Raw Accelerometer Data

**DOI:** 10.64898/2026.05.28.26354364

**Authors:** Pin-Wei Chen, Christopher Cielo, Olivia Walch, Morgan McDonald, Peter X.K. Song, Cathy Goldstein, Jennette P. Moreno, Erica C. Jansen, Jonathan A. Mitchell

## Abstract

**Introduction:** Actigraphy sleep-wake classification methods increasingly seek to leverage raw acceleration data and machine-learning-based classification, but performance evaluation in pediatrics is limited. We trained machine-learning models using pediatric data and compared their sleep-wake classification performance with existing algorithms for children.

**Methods:** Sixty-five children (46% female, ages 5.3–17.7 years) completed in-lab overnight polysomnography and wore a GENEActiv device on their non-dominant wrist. The acceleration data were converted into 30-second epochs and aligned with physician-scored sleep-wake data from electroencephalography. Seven machine-learning models were trained using leave-one-subject-out cross-validation. Epoch-by-epoch analyses generated performance metrics (e.g., balanced accuracy [BA]) and discrepancy analyses provided overall sleep duration bias estimates. The combination of highest performance and least bias was used to rank using Euclidean distance scores - where a lower score represents closer to perfect performance and zero bias. For benchmarking, we included GGIR sleep scoring algorithms and an adult trained random forest classifier.

**Results:** Overall, 560.1 hours of polysomnography and actigraphy data were collected (74.4% of epochs were scored as sleep). The pediatric-trained local-global long-short term memory (LSTM) classifier had the most optimal epoch-by-epoch performance (e.g., BA=0.85, sensitivity=0.88, specificity=0.83, ROC-AUC=0.95, and Cohen’s kappa=0.67). These metrics exceeded that of an adult-trained random forest classifier and GGIR-based algorithms. Discrepancy analyses revealed that overall sleep duration was underestimated by an average of 25 minutes using the LSTM classifier with no proportional bias.

**Conclusion:** We trained seven pediatric sleep-wake classifiers that had strong ability to detect sleep and wake, with the LSTM classifier being most optimal.

## Introduction

Sleep is an evolutionarily conserved circadian-regulated process that has important implications for health and development in childhood^1-8^. Sleep assessment is therefore needed in pediatric research and healthcare, and actigraphy is a prominent methodology used for non-intrusive assessment over multiple nights in the home environment^9^. Increasingly, actigraphy methods leverage raw sensor data and open-source sleep scoring algorithms to generate sleep metrics. This produces device-independent sleep estimates, and the data can be re-processed in the future as more optimal scoring methods become available. However, there are a limited number of computational tools to process raw sensor data for sleep estimation, and it may not be known if the methods used are suitable for sleep estimation in children whose sleep behavior can differ from adults^10-12^.

GGIR is an open-source R package that serves as an end-to-end computational tool for raw acceleration data processing, which includes pre-processing, non-wear detection, sleep-wake scoring, and sleep period detection steps^13^. We and others have documented the sleep-wake scoring performance of three GGIR-based algorithms in children^14-16^. Interestingly, the Sadeh algorithm^17^, which was originally developed for use in children, and has been adapted and integrated for use in the GGIR workflow, was found to have the least optimal performance in two pediatric validation efforts^14, 15^. It is therefore possible that a more optimal pediatric sleep-wake scoring algorithm could be developed for raw sensor data processing, like GGIR. Indeed, machine learning methods are increasingly being used to develop sleep-wake classifiers using biometric features, and such methods may help to improve actigraphy methods for sleep assessment in children.

Machine learning methods are well suited at handling high-resolution time series sensor data, such as accelerometer data, and to incorporate potential non-linear relationships and interactions between multiple motion features. Further, these models are adaptable, and their performance can potentially improve when more training data become available. Prior studies have assessed the sleep-wake detection performance using a variety of machine learning methods, and models using deep learning architectures tend to produce higher performing sleep-wake classifiers^10, 18-22^. However, most prior studies tended to focus on adults and did not systematically assess each machine learning taxonomy. For example, in adults, Walsh et al. found that sleep-wake classifiers developed using neural network, logistic regression, and random forest models performed strongly^10^ and Shi et al. showed convolution neural network models performed strongly^18^. In children, Weaver et al. found that a local-global long short-term memory (LSTM) classifier had the most optimal sleep-wake classification performance compared to traditional logistic regression and random forest models^19^.

Building on prior research, we aimed to determine the sleep-wake detection performance of pediatric-trained machine learning classifiers. This validation study used features from raw accelerometer data and labels of physician scored sleep-wake states derived from electroencephalography. We trained seven machine learning models, assessed their performance at detecting sleep-wake in the lab setting, and benchmarked against GGIR-based sleep-wake scoring algorithms and an adult-trained machine learning sleep-wake classifier. The selection of the seven machine learning algorithms was based on prior sleep-wake classifiers^10, 19-22^, with the aim to include one model per machine learning taxonomy as there are benefits to different methods. From prior research, we expected that random forest, neural network, logistic regression, and local-global LSTM would have the most optimal sleep-wake classification performance.

## Methods

### Sample

We enrolled typically developing children, aged 5-17 years, with no prior diagnosis of a sleep disorder, who were completing an in-lab overnight sleep study at the Children’s Hospital of Philadelphia’s Sleep Laboratory. The electronic health records were reviewed for patients scheduled for polysomnography to assess eligibility (within age range, no documentation of a prior sleep disorder diagnosis, no documentation of a prescribed medication that could impact sleep, and no documentation of a chronic medical condition requiring ongoing care). Passing this medical record screening step, we asked caregivers to complete a screening questionnaire to confirm eligibility. The in-lab data were collected between 2022-2026. Most of the participants enrolled were clinically referred for polysomnography for evaluation of obstructive sleep apnea (OSA; the most common reason for referral, with an expectation that approximately half would receive an OSA diagnosis). We also enrolled children who were completing an overnight sleep study as part of another research study. A study coordinator provided the actigraphy device, and the sleep lab personnel completed the polysomnography procedures. Participants provided their assent and a parent/legal guardian accompanying their child provided consent. The study was approved by the CHOP Institutional Review Board (IRB, 19-016592).

### Polysomnography Procedures

Overnight, in-lab sleep studies were conducted at CHOP’s Sleep Laboratory following the American Academy of Sleep Medicine (AASM) pediatric guidelines^23^. Participants arrived with a parent or legal guardian in the early evening (typically around 7:00 PM) and departed soon after waking the next day (typically around 6:30 AM). Data collection utilized a Polysmith polysomnography system (Nihon Kohden, Irvine, CA) and included the following measurements: electroencephalography, bilateral electrooculograms, submental and tibial electromyograms, respiratory inductance plethysmography, electrocardiogram, pulse oximetry, infrared capnometry, a three-pronged thermistor, and nasal pressure. Continuous audio and video were captured with an infrared camera and microphone to allow a technician to monitor participants in the darkened room. Initial review of polysomnography data was performed by a sleep technologist, followed by scoring by Dr. Cielo, a CHOP board-certified pediatric sleep medicine physician. Scoring adhered to the AASM Manual for the Scoring of Sleep and Associated Events^23^, with sleep or wake states classified in 30-second epochs.

### Actigraphy Procedures

During the in-lab sleep assessment, each participant wore a GENEActiv actigraphy device (Activinsights, Cambridge, UK) on their non-dominant wrist and the time of placement was documented. The devices were configured to record tri-axial acceleration at a 50 Hz sampling rate. After the overnight study concluded in the morning, the actigraphy device was removed, and the raw x, y, and z axis acceleration data were downloaded and stored for analyses.

### Machine Learning Training and Testing

#### Machine Learning Method Selection

We selected methods that cover each machine learning taxonomy. Six classical procedures and one deep learning machinery were trained including 1) a linear logistic regression model, 2) perceptron models (single hidden layer neural network), 3) instance-based K nearest neighbor, 4) tree-based models (decision trees), 5) kernel-based models (support vector machine), 6) an ensemble machine learning model (random forest and extreme gradient boosting), and 7) a deep learning model (LSTM).

Machinery architecture and associated hyperparameter search grids were determined based on established optimality objectives algorithms and software with demonstrated efficacy in sleep-wake detection among adult and pediatric populations. For the deep learning approach, we adapted the LSTM architecture proposed by Weaver et al.^19^, a unique design that integrates bidirectional local and global LSTM, and was shown to have strong performance in a pediatric sample. Regardless of the underlying architecture, all models were subjected to standardized preprocessing pipelines and nested cross-validation procedures, with hyperparameters tuning optimized via rigorous grid search.

#### Software and Computational Environment

All data processing was done in R (version 4.4.0) with Tidyverse (version 2.0.0)^24^, and data.table packages (version 1.17.8)^25^. The six classical machine learning methods were processed with Tidymodel (version 1.4.1)^26^. The LSTM was processed in Python (version 3.10.4) and PyTorch (version 2.1.0). The machine learning procedures used the following packages: ranger package (version 0.17.0)^27^for Random Forest, kknn package (version 1.4.1)^28^ for k-Nearest Neighbors, glmnet package (version 4.1.10)^29^ for Logistic Regression, xgboost (version 1.7.11.1)^30^ for Extreme Gradient Boost, rpart package (version 4.1.23)^31^ for Decision Tree, and nnet package (version 7.3-19)^32^ for Neural Network. We utilized generative artificial intelligence in the process of code development to screen for coding bugs and generate fixes with human supervision.

#### Data Pre-Processing

First, periods when the standard deviation of the vector magnitude was <0.02 gravitational unit were flagged, as this is indicative of non-wear. These periods were identified towards the end of the sleep test for 12 participants and were removed when there was a corresponding wake time polysomnography label. Second, missing values, which can occur from device malfunction, were imputed using k-nearest neighbors at the level of feature space. Imputation was necessary because most machine learning algorithms cannot function with gaps in the data, and simply discarding incomplete data often leads to biased or less accurate models. Third, all features (except frequency amplitudes) were z-score normalized to minimize the potential influence of baseline values and outliers on the model. Fourth, highly correlated features (correlation > 0.9) were removed to prevent overfitting on similar features.

#### Feature Generation

Raw accelerometer data from the x, y, z axes, and vector magnitude were calculated into non-overlapping 30-second epoch features and aligned with the sleep or wake labels from the physician-scored polysomnography data. The features are a series of calculations that describe the 30-second wave form of x, y, z, and vector magnitude. The vector magnitude (i.e., magnitude) was calculated to capture the overall intensity of movement independent of device orientation:

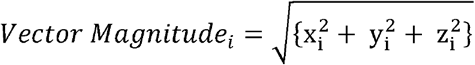

where *x*_*i*_,*y*_*i*_, and *z*_*i*_ represent the 3-dimensional acceleration samples at time index *i*. The resulting feature set comprised statistical time-domain metrics and frequency-domain characteristics for each axis (*x,y,z*) and the magnitude. The time-domain metrics are average, standard deviation, range, median, interquartile range, mode, and zero cross rate (excluding magnitude). The frequency domain included dominant frequency and spectral energy. These features are used in the prior literature of actigraphy-based sleep-wake or motion detection algorithms^22^. Thirty-five features were derived (Supplementary Table 1). For LSTM, feature generation followed prior research using spectral density from an overlapping moving window with a 1 Hz to 30 Hz sampling frequency per 30-second epoch. The spectral density features should not be normalized in the preprocessing steps because it decreased the accuracy based on our prior validation metrics based on our internal testing.

#### Cross-validation

A nested cross-validation was used to maximize the determination of optimal model tuning and performance evaluation (Supplementary Figure 1). The outer loop employed leave-one-subject-out cross-validation (LOSOCV), while the inner loop performed hyperparameter tuning via 5-fold cross-validation. The outer loop determines the performance of each model and uses all subjects’ data as training data except one as testing data, rotating across all subjects. Within each cycle of LOSOCV, we performed an inner loop cross-validation where 80% of the data were training data and 20% data were testing data and each cycle rotated across the data splits to find the optimal hyperparameters maximized in the area under the receiver-operating characteristic curve (ROC-AUC). The weighted cross entropy loss was calculated during each pass of the LSTM data in the inner loop with a maximum of 100 passes. An early stop was implemented at the occurrence of 5 consecutive identical scores to prevent overfitting.

#### Class Imbalance

During typical overnight sleep periods, there are more sleep epochs than wake epochs. Left unaccounted for, the machine learning model will be biased toward predicting sleep over wake. To counter this bias, we used the Synthetic Minority Oversampling Technique (SMOTE)^33^. SMOTE works by finding k-nearest neighbors of minority samples and generating new points along the lines connecting them in the feature space, adding variety and improving model generalization. This process was applied to the training data set and not the testing data within the cross-validation. SMOTE was applied to all models except the LSTM. Instead, based on the prior research from Weaver et al., weighted cross-entropy loss was used so that it can better account for the imbalance in the temporal dimension and the deep learning structure in the LSTM.

#### Existing Sleep-Wake Algorithms for Benchmarking

To benchmark our pediatric-trained models, we compared them against existing methods: GGIR-based sleep algorithms and an adult-trained random forest sleep-wake classifier. For the former, GGIR version 3.3-4^13^ was used to generate sleep-wake estimates using the van Hees (GGIR-vH), Cole-Kripke (GGIR-CK), and Sadeh (GGIR-S) algorithms using the same methods we reported in a prior study. For the latter, an adult-data trained random forest classifier^10^ was used to generate sleep-wake estimates. This model was trained from 31 adult subjects (age: 19-55 years, 68% female) and tested on 188 subjects from Multi-Ethnic Study of Atherosclerosis (MESA) dataset (age: 56-89 years, 48% female)^10^.

### Statistical Analyses

To describe the sample, we used means (standard deviations) and medians (interquartile ranges) for continuous variable, and frequencies (percentages) for categorical variables. To determine the performance of each sleep-wake classifier, we completed epoch-by-epoch and discrepancy analyses using open-source R package from Menghini et al.^34^ For the epoch-by-epoch analysis, the accelerometer data were aligned into 30-second epochs against the labeled polysomnography sleep dataset. Epoch-level sleep-wake detection performance was assessed using sensitivity, specificity, F1 scores (harmonic mean of sensitivity and precision), balanced accuracy (BA; arithmetic mean of sensitivity and specificity), receiver operated area under the curve (ROC-AUC), Cohen’s Kappa agreement, and Matthews correlation coefficient (MCC)^35^. Sleep epochs were labeled as positive classes and wake epochs were labeled as negative classes. For reference purposes, we included the performance metrics for a naïve method that predicted all epochs as sleep or all epochs as wake. For the discrepancy analysis (i.e., night-to-night comparison), we compared the night-level sleep data estimated with Bland-Altman plots generated. Log transformation was used when data were skewed. To rank the algorithms, we calculated the average Euclidean distances from perfect performance and zero overall bias, with weighting applied to correct for time spent awake during the sleep period, using the following formula:

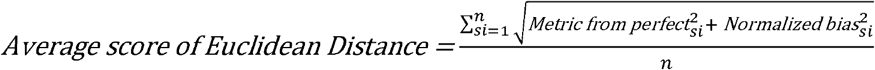

Where *Normalized Bias*_*si*_ *= bias*_*si*_ / *max*_*global*_ and n = is the total number of subject and *si* is the subject index.

## Results

Sixty-five children completed the overnight sleep test and provided actigraphy data (Table 1). The average age of the sample was 11.7 (SD=3.0) years and ranged from 5.3 to 17.7 years. The sample was 46% female, 22% White, 58.6% Black, and 86.2% were clinically referred for suspected OSA. From polysomnography, most did not have OSA (55%) but 26%, 12%, and 6% were diagnosed with mild, moderate, and severe OSA, respectively. Overall, 560.1 hours of overnight polysomnography data were collected, with 74% of epochs scored as sleep (Table 1). The average polysomnography determined sleep duration was 6.9 (SD=1.3) hours (Table 1).

**Table 1.** Characteristics of the study sample.

| **Characteristic** | **All Participants**<br>**(N=65)** |
| --- | --- |
| Age, mean (SD), years | 11.7 (3.0) |
| Female, N (%) | 30 (46.0) |
| Black, N (%) | 38 (58.6) |
| Hispanic, N (%) | 3 (4.6) |
| White, N (%) | 14 (22.0) |
| Asian, N (%) | 1 (1.5) |
| More than One Race, N (%) | 5 (7.7) |
| Unknown or Other, N (%) | 4 (6.1) |
| BMI, mean (SD), Percentile | 83.2 (24.1) |
| Clinically Referred, N (%) | 56 (86.2) |
| Research Participants, N (%) | 9 (13.8) |
| AHI, median (IQR), index | 1.3 (0.7, 4.8) |
| Arousal Index, median (IQR), index | 10.5 (7.3, 14.4) |
| No OSA, N (%) | 36 (55.3) |
| Mild OSA, N (%) | 17 (26.2) |
| Moderate OSA, N (%) | 8 (12.3) |
| Severe OSA, N (%) | 4 (6.2) |
| Sleep Duration, mean (SD), hours | 6.9 (1.3) |
| Wake After Sleep Onset, mean (SD), minutes | 59.6 (47.5) |
| Sleep Latency, mean (SD), minutes | 44.1 (36.3) |

AHI, Apnea-hypopnea index; BMI, body mass index; OSA, obstructive sleep apnea

### Preprocessing

For the 12 participants who had periods of non-wear (standard deviation of the vector magnitude was <0.02 gravitational units and polysomnography wake label), an average of 124.4 minutes were removed with standard deviation of 79.7 minutes (minimum 5 minutes and maximum 298 minutes removed). Imputation and feature removal were done independently in each leave-one subject-out cross-validation. During the feature imputation step, two data points of the standard deviation and zero crossing rate among x, y, z and vector magnitude columns were imputed (approximately 0.28% of the total training data). During the feature removal steps, fifteen features were commonly identified as highly correlated (greater than 90%) and were removed. One feature, the magnitude of the dominant frequency, was removed due to zero variance. Four features were removed inconsistently (i.e., 2, 3, 22 or 43 times) during the cross-validations. The details of the number in feature removal can be found in Supplementary Table 2.

### Epoch-by-Epoch Analyses

Of the seven pediatric-trained machine learning models (Table 2), LSTM had the highest sensitivity (0.88), specificity (0.83), F1 score (0.91), balanced accuracy (0.85), ROC-AUC (0.95), Cohen’s Kappa (0.67), and MCC (0.68). The neural network, random forest, and logistic regression classifiers performed strongly, but to a lesser extent than the LSTM classifier (e.g., neural network had an ROC-AUC of 0.84). The decision tree, extreme gradient boosting, and k nearest neighbor classifiers had the lowest sleep-wake detection performance metrics (e.g., k nearest neighbor had an ROC-AUC of 0.75). Compared to GGIR-based sleep scoring algorithms, the LSTM classifier outperformed GGIR-vH, GGIR-CK, and GGIR-S (e.g., GGIR-vH had balanced accuracy of 0.78). The LSTM classifier also outperformed an adult trained random forest classifier (ROC-AUC of 0.85).

The sleep-wake detection performance of the pediatric trained classifiers were generally consistent for male and female children, across the sample age range, and for those with and without OSA (Supplementary Table 3). However, some differences were detected for sensitivity and specificity performance metrics for certain classifiers (Supplementary Table 3), and after correcting for multiple testing using family wise error rate using post-hoc Bonferroni correction, we only observed that older age was associated with higher sensitivity for the logistic regression classifier.

**Table 2.**
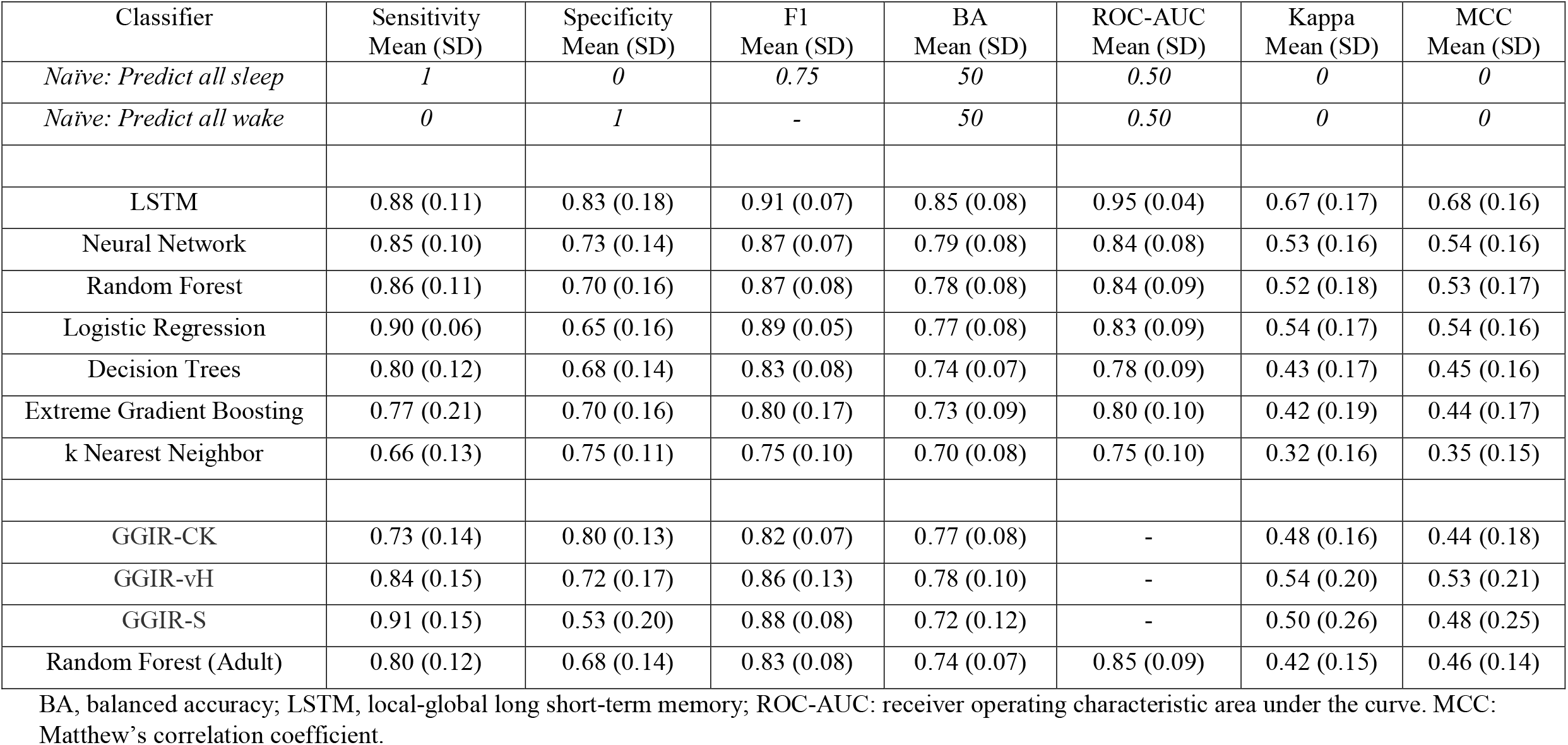
Sleep-wake detection performance of classifiers against EEG determined sleep.

| Classifier | Sensitivity<br>Mean (SD) | Specificity<br>Mean (SD) | F1<br>Mean (SD) | BA<br>Mean (SD) | ROC-AUC<br>Mean (SD) | Kappa<br>Mean (SD) | MCC<br>Mean (SD) |
| --- | --- | --- | --- | --- | --- | --- | --- |
| *Naïve: Predict all sleep* | *1* | *0* | *0.75* | *50* | *0.50* | *0* | *0* |
| *Naïve: Predict all wake* | *0* | *1* | *-* | *50* | *0.50* | *0* | *0* |
| LSTM | 0.88 (0.11) | 0.83 (0.18) | 0.91 (0.07) | 0.85 (0.08) | 0.95 (0.04) | 0.67 (0.17) | 0.68 (0.16) |
| Neural Network | 0.85 (0.10) | 0.73 (0.14) | 0.87 (0.07) | 0.79 (0.08) | 0.84 (0.08) | 0.53 (0.16) | 0.54 (0.16) |
| Random Forest | 0.86 (0.11) | 0.70 (0.16) | 0.87 (0.08) | 0.78 (0.08) | 0.84 (0.09) | 0.52 (0.18) | 0.53 (0.17) |
| Logistic Regression | 0.90 (0.06) | 0.65 (0.16) | 0.89 (0.05) | 0.77 (0.08) | 0.83 (0.09) | 0.54 (0.17) | 0.54 (0.16) |
| Decision Trees | 0.80 (0.12) | 0.68 (0.14) | 0.83 (0.08) | 0.74 (0.07) | 0.78 (0.09) | 0.43 (0.17) | 0.45 (0.16) |
| Extreme Gradient Boosting | 0.77 (0.21) | 0.70 (0.16) | 0.80 (0.17) | 0.73 (0.09) | 0.80 (0.10) | 0.42 (0.19) | 0.44 (0.17) |
| k Nearest Neighbor | 0.66 (0.13) | 0.75 (0.11) | 0.75 (0.10) | 0.70 (0.08) | 0.75 (0.10) | 0.32 (0.16) | 0.35 (0.15) |
| GGIR-CK | 0.73 (0.14) | 0.80 (0.13) | 0.82 (0.07) | 0.77 (0.08) | - | 0.48 (0.16) | 0.44 (0.18) |
| GGIR-vH | 0.84 (0.15) | 0.72 (0.17) | 0.86 (0.13) | 0.78 (0.10) | - | 0.54 (0.20) | 0.53 (0.21) |
| GGIR-S | 0.91 (0.15) | 0.53 (0.20) | 0.88 (0.08) | 0.72 (0.12) | - | 0.50 (0.26) | 0.48 (0.25) |
| Random Forest (Adult) | 0.80 (0.12) | 0.68 (0.14) | 0.83 (0.08) | 0.74 (0.07) | 0.85 (0.09) | 0.42 (0.15) | 0.46 (0.14) |

BA, balanced accuracy; LSTM, local-global long short-term memory; ROC-AUC: receiver operating characteristic area under the curve. MCC: Matthew's correlation coefficient.

### Discrepancy Analyses

The discrepancy analyses are presented in Table 3, and corresponding proportional bias plots are in Supplementary Figures 2-9. Of all seven pediatric-trained classifiers, the one with the least biased estimate of sleep duration was logistic regression, with an average overestimation of 5 minutes (Table 3). However, the bias was proportional with a shift from overestimation to underestimation as sleep duration increased (Supplementary Figure 6). The LSTM classifier was the only pediatric-trained classifier that did not exhibit proportional bias. It underestimated sleep duration by an average of 30 minutes (Table 4). Regarding the benchmark algorithms, only GGIR-vH and GGIR-S did not have proportional bias; the former underestimated sleep duration by an average of 33 minutes and the latter overestimated sleep duration by 27 minutes (Table 3)

**Table 3.**
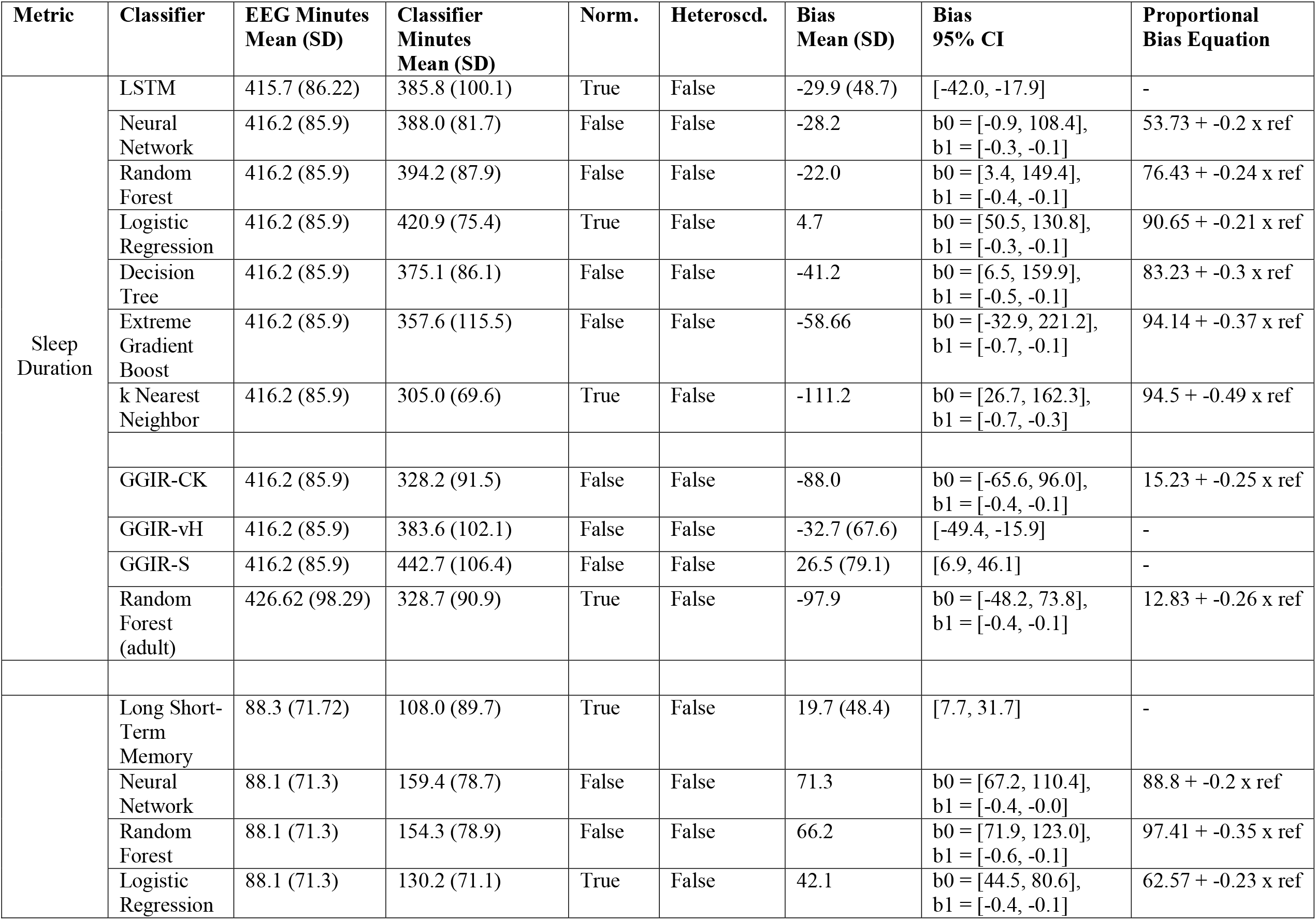
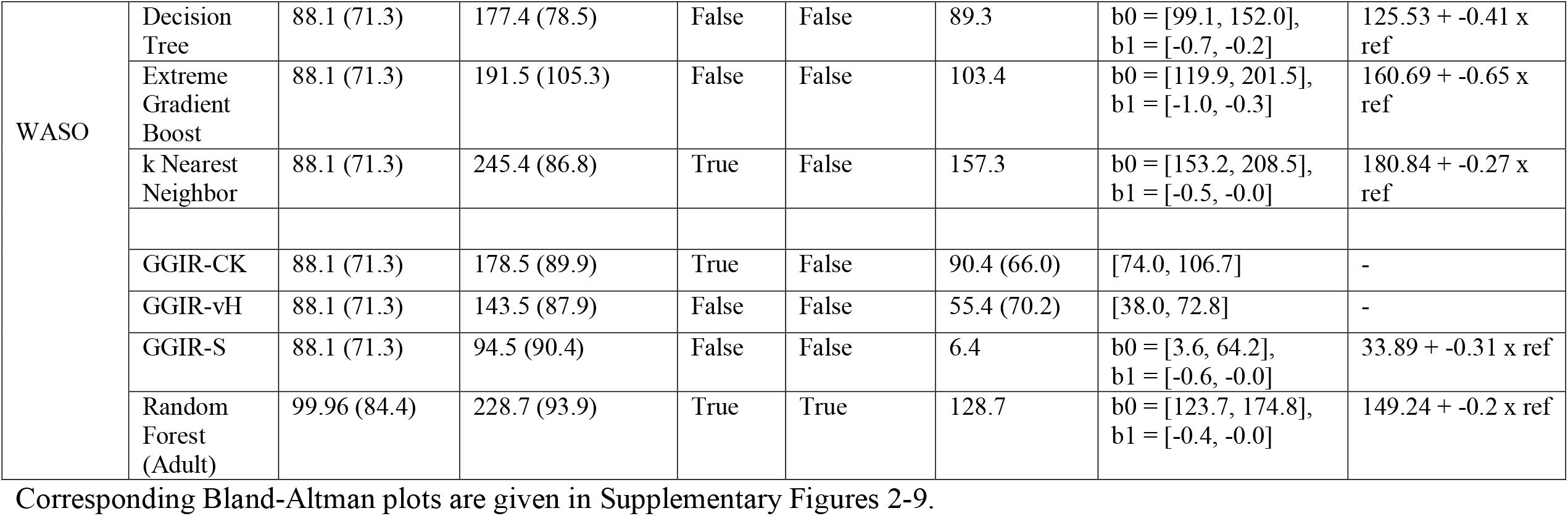
Discrepancy analysis results and detection of sleep duration and WASO bias.

Regarding WASO, the pediatric-trained classifier with the least bias was the LSTM classifier, with an overestimation of 20 minutes and no evidence of proportional bias (Table 3). All other pediatric-trained classifiers had a larger degree of WASO overestimation with proportional bias present (Table 4). Regarding the benchmark algorithms, GGIR-S had the least bias with an overestimation of 4 minutes, but proportional bias was present (Table 3).

### Ranking Score

Scatterplots for epoch-by-epoch performance metrics and overall sleep duration are presented in Figure 1, with Euclidian distance composite scores provided to rank classifier performance. Using balanced accuracy, the LSTM (score=0.18), neural network (score=0.24), and logistic regression (0.24) were the top three pediatric-trained classifiers (Figure 1). Regarding the benchmarking algorithms, GGIR-vH had composite scores of 0.25 and the adult trained random forest model had a score of 0.33 for balanced accuracy (Figure 1). The overall ranking for each performance metric is given in Figure 2. LSTM ranked 1^st^ or 2^nd^ for all metrics. The corresponding figures for WASO are given in Supplementary Figure 10 and 11; the LSTM classifier ranked 1^st^ for all metrics regarding WASO.

**Figure 1.**
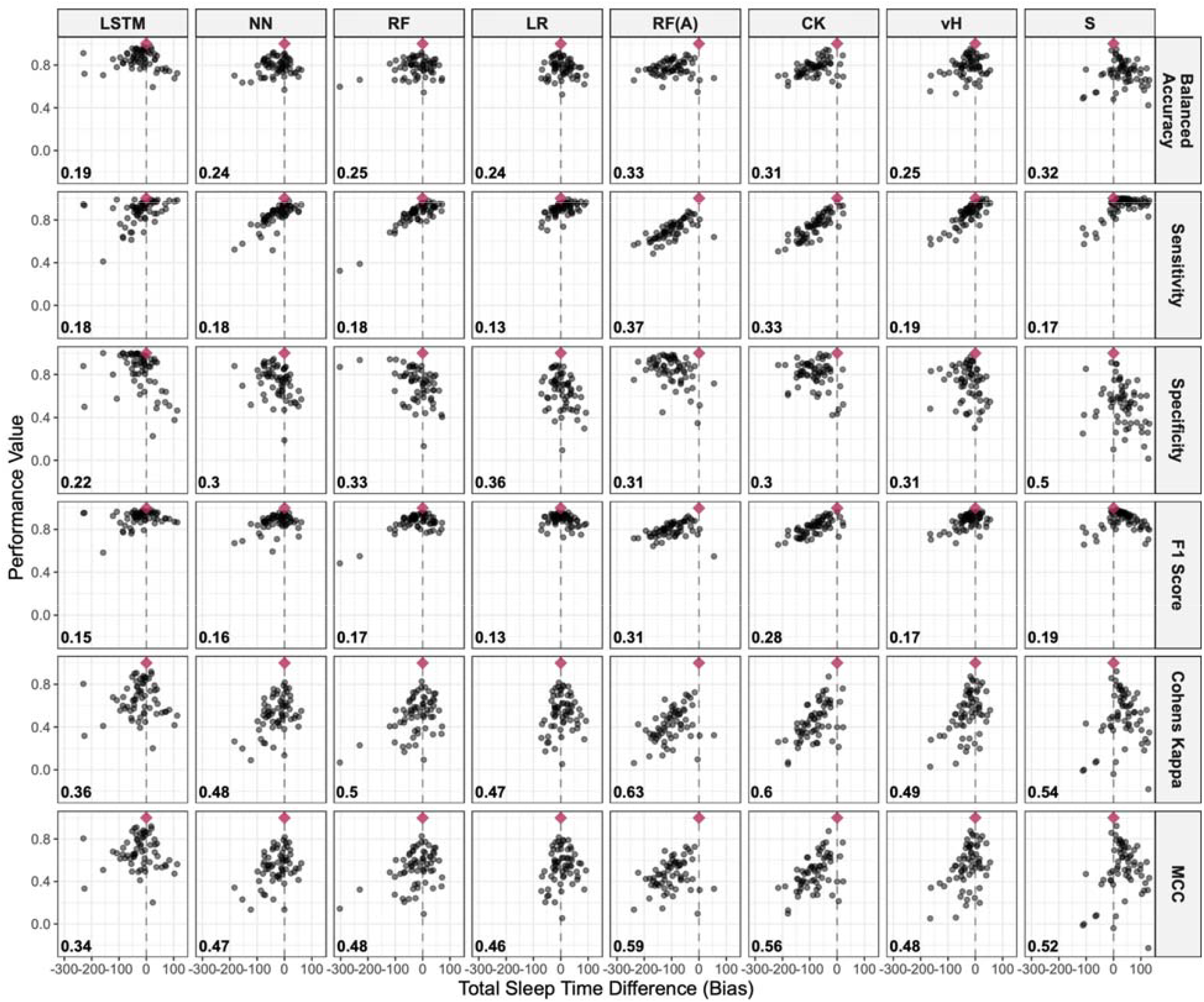
Scatter plot of total sleep time bias versus performance metrics for the top 4 performing pediatric trained classifiers and the benchmarking methods. The red diamond indicates a perfect score (0 minutes of bias and a metric score of 1). The dashed line represents 0 minutes of bias. Tighter clustering indicates higher model reliability, while proximity to the red diamond signifies better overall model performance. The Euclidian distance scores from perfect performance and zero bias are given in the lower left corner. LSTM = Long Short-Term Memory, NN = Neural Network, RF = Random Forest, LR = Logistic Regression, RF(A) = Random Forest (Adult), CK = GGIR-adapted Cole-Kripke, vH = GGIR-adapted van Hees, S = GGIR-adapted Sadeh.

**Figure 2.**
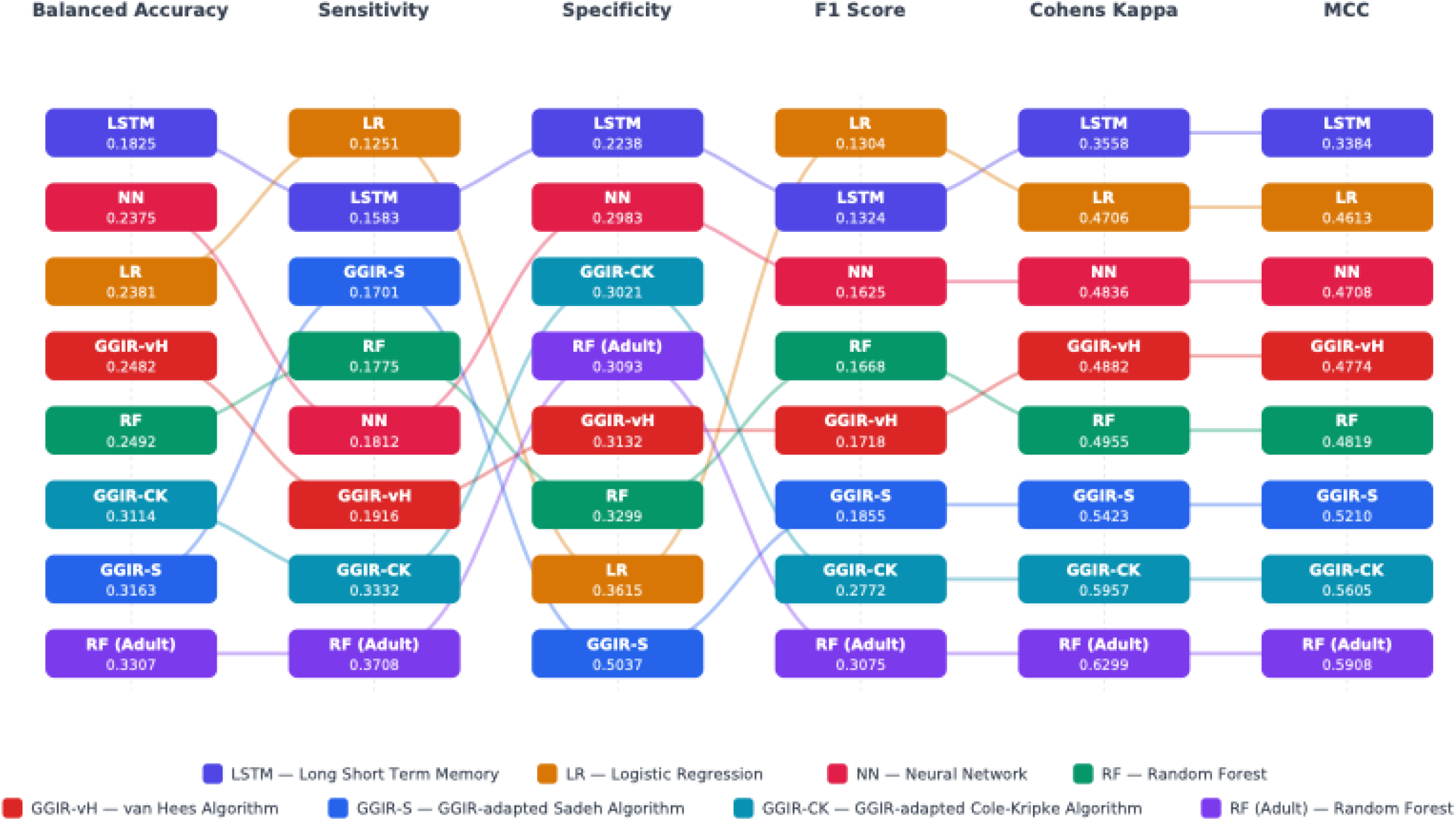
Sankey diagram of Euclidean Composite score for the sleep duration bias and balanced accuracy score of the top machine learning and benchmark references.

## Discussion

Actigraphy is a key wearable method to estimate sleep over multiple nights in pediatric research and healthcare. The collection and availability of raw, unprocessed sensor data provide opportunities to develop novel sleep-wake scoring methods using machine learning. Leveraging pediatric data, we trained seven machine learning models, covering each machine learning taxonomy, and compared model derived sleep-wake classifications with physician labeled sleep-wake data from polysomnography in 30 second epochs. We observed that the LSTM classifier was most optimal based on epoch-by-epoch and discrepancy analyses. Benchmarking steps revealed that the performance of this classifier exceeded that of an existing random forest classifier trained with adult data, and GGIR-based sleep scoring algorithms. With independent verification, these pediatric trained sleep–wake classifiers could enhance the scoring of wearable sensor data for sleep estimation in children, especially the local-global LSTM classifier.

Few prior studies have used machine learning methods for pediatric sleep classification using actigraphy and polysomnography data. Migovich et al.^36^ evaluated a decision tree classifier for three-stage sleep classification (awake, Non-REM, or REM) in seven children with Rett syndrome (aged 4–16 years) using a wrist-worn Empatica E4. Their accelerometer-only model achieved 0.94 overall accuracy for NREM detection but performed poorly in classifying wake (0.49 accuracy) and REM sleep (0.29 accuracy) states^36^. Note, this study only reported overall accuracy which can be biased by the class imbalance between REM, non-REM, and wake time. Tilmanne et al.^37^ used data from ankle-placed actigraphy in 354 infants and applied various algorithms, including decision trees, multilayer perceptrons (neural network), and the Sadeh and Sazonov heuristics. Across all configurations, sleep detection sensitivity was high (0.81-0.95), whereas specificity for detecting wakefulness was notably lower (0.39-0.69); the best-performing model was a decision tree classifier that achieved 0.92 sensitivity, 0.69 specificity, and 0.80 balanced accuracy^37^. More recently, Weaver et al.^19^ evaluated local-global LSTM, logistic regression, and random forest classifiers in a cohort of 238 children (aged 5–12 years) with suspected sleep disruptions. Participants wore an ActiGraph GT9X on the non-dominant wrist concurrently with consumer wearables during an overnight in-lab sleep test^19^. For binary sleep-wake classification using actigraphy alone, the LSTM classifier had the most optimal performance, achieving a sensitivity of 0.95-0.96 and specificity of 0.85-0.90^19^. Our results support the high performance of the deep learning LSTM classifier for sleep-wake detection in children using accelerometry features.

Comparisons can also be drawn from adult-based actigraphy and polysomnography studies. Palotti et al.^11^ compared ten models — four traditional ML (Extra Trees, Logistic Regression, Linear SVM, Perceptron) and six deep learning (LSTM and Convolution Neural Network (CNN) at 20, 50, and 100 hidden units) — on 1,817 adults using data from wrist-placed Actiwatch Spectrum devices. The best-performing model was LSTM with 100 hidden units, which achieved sensitivity of 0.91, specificity of 0.70, balanced accuracy of 0.81, and F1 score of 0.86^11^. Sundararajan et al.^20^ evaluated four algorithms (Sadeh, Cole-Kripke, GGIR-vH, and random forest) for sleep-wake classification on 134 adults (aged 20–72 years; 64 healthy sleepers and 70 with sleep disorders) using a wrist-worn GENEActiv accelerometer. Notably, this study used nested cross-validation, and reported sleep-wake detection performance via F1 score and Cohen’s kappa. The best-performing model was the random forest classifier, which achieved an F1 score of 0.76 and kappa of 0.52^20^. Patterson et al.^12^ tested 8 different algorithms (Webster, Sadeh, Cole-Kripke, Oakley, Sazonov, CNN, LSTM, GGIR-vH) in adults (MESA cohort, 54-93 years of age); the best-performing model was LSTM 100 hidden unit sleep-wake classifier with 0.79 sensitivity, 0.59 specificity, and 0.69 balanced accuracy^12^.

Walch et al.^10^ tested four classifiers (logistic regression, k-nearest neighbors, random forest, and neural network) using Apple Watch data on 31 healthy adults with concurrent polysomnography and validated with external 188 older adult cohort from MESA dataset. Using motion only features, most classifiers had area under the ROC curve (ROC-AUC) that exceeded 0.8^10^; the neural network classifier had an ROC-AUC of 0.82, random forest ROC-AUC was 0.81, and logistic regression ROC-AUC was 0.82^10^. We included the adult trained random forest classifier from Walch et al. for benchmarking purposes and replicated the same level of sleep-wake detection performance in children, with an AUC of 0.85. However, this performance was below that of four pediatric trained classifiers in our study, especially the LSTM classifier.

We also benchmarked against GGIR based sleep scoring algorithms. In a prior study, we reported the sleep-wake scoring performances of GGIR-based sleep scoring algorithms in a sample of 30 children^14^. Using the same methods, but in an expanded sample of 65 children, we replicated the original performance metrics with minor modifications, whereby GGIR-vH and GGIR-CK continued to have high sensitivity and moderate specificity, and GGIR-S continued to have very high sensitivity and lower specificity. These expanded results show that the sleep-wake scoring performance of GGIR-based sleep algorithms are stable and therefore strong for benchmarking purposes. We found that most of our pediatric trained classifiers were comparable or exceeded the sleep-wake detection performance of GGIR-vH and GGIR-CK, and were superior to GGIR-S, especially the LSTM classifier. This is notable, since the Sadeh algorithm was developed in 1994 for specific use in children^17^. Our results indicate that caution should be taken when using the GGIR version of the Sadeh algorithm in children as there are more optimal sleep-wake detection methods available.

The top-performing architecture in our evaluation was the local-global LSTM, adapted from the framework established by Weaver et al.^19^. We hypothesize that this model outperformed alternative architectures—including standard LSTMs—due to its dual-stream processing capability. Specifically, the model captures fine-grained regional dependencies within frequency density patterns while simultaneously leveraging the predictable pattern of the global temporal structure of sleep. This allows the model to account for the physiological tendency of sleep stages to persist in stable clusters rather than frequently transitioning between disparate states.

Our study includes strengths and limitations. We comprehensively evaluated classic machine learning ensembles and benchmarked against open-source GGIR-based sleep-wake algorithms and an adult-trained sleep-wake classifier. We provided multiple epoch-by-epoch performance metrics to allow for comparisons with prior studies. We proposed a new composite Euclidian distance score to help rank sleep-wake scoring methods that considers performance metrics and overall night-level bias. Our results are most generalizable to typically developing children and children being evaluated for OSA. Critically, we found no major age, sex or OSA status differences in classifier performance, but larger scale studies are needed to fully assess performance across demographic and clinical groups (i.e., we are currently limited by a small number of children with severe OSA in our study and cannot claim generalizability of the model performance to severe OSA). Additional studies should evaluate cohorts of children for whom these findings may not be generalizable, including those with neuromuscular disease or Down syndrome. We did not have an independent testing data set; collaborating on multi-site efforts or multi-data from the same subject would address this limitation. Finally, the models developed require technical skills to apply and are not yet integrated for use in cohort studies. We have developed a cloud-based platform to process raw acceleration data for sleep estimation^38^, and we plan to integrate our models (especially the LSTM) into this platform in the future.

In conclusion, we developed pediatric sleep-wake classifiers demonstrating robust sleep detection and wake detection. Through epoch-by-epoch and discrepancy analyses, the LSTM classifier emerged as the optimal model, surpassing GGIR based sleep scoring algorithms, and an adult-trained random forest classifier. Future studies utilizing larger training and validation datasets may further minimize variability and enhance the precision of accelerometry-based pediatric sleep-wake classification. Given the high performance of the LSTM method, other deep learning model architectures should be considered in the future.

## Data Availability

All data produced in the present study are available upon reasonable request to the authors.

## Acknowledgments

The reported research in this publication was supported by the Eunice Kennedy Shriver National Institute of Child Health & Human Development of the National Institutes of Health under Award Number R01HD100421. The content is solely the responsibility of the authors and does not necessarily represent the official views of the National Institutes of Health.

