## Supplementary File for "The Sleep-Wake Classification Performance of Pediatric-Trained Machine Learning Algorithms for Raw Accelerometer Data"

Supplementary Table 1. Thirty-Five Features generated

| Time Domain | Statistical descriptors were computed for each axis and the magnitude. |
| --- | --- |
| Average | The arithmetic average of the signal values. |
| Standard Deviation | A measure of the dispersion of the signal values. |
| Range | The difference between the maximum and minimum amplitude. |
| Median | The middle value separating the higher half from the lower half of the data sample. |
| Interquartile Range | A measure of statistical dispersion, calculated as the difference between the 75th percentile and the 25th percentile. |
| Mode | The most frequently occurring value in the signal. To calculate the mode for continuous accelerometer data, values were first discretized by rounding to two decimal places. |
| Zero Cross Rate | To measure signal oscillation, we calculated the rate at which the signal crosses its own mean. |
| Frequency Domain | A Fast Fourier Transform (FFT) was applied to transform the time-series data into the frequency domain. Features were derived from the magnitude of the FFT coefficients up to the Nyquist frequency (25 Hz). |
| Dominant Frequency | The frequency component associated with the maximum spectral magnitude. |
| Spectral Energy | The sum of the squared magnitudes of the frequency components, representing the signal's total strength in the frequency domain. |
| Power spectrum density value (LSTM) | The data were up-sampled to 100Hz then transformed into 1-second overlapped moving segments. Then we used FFT to create the power spectrum then calculate the magnitude of the complex FFT coefficients. Then the average of each frequency was calculated. |
| Raw accelerometer data from the x, y, and z axes were segmented into non-overlapping 30-second epochs. For every epoch, a Signal Vector Magnitude (SVM) was calculated to capture the overall intensity of movement independent of device orientation:$SVM_{i}=\sqrt{\left\{ x_{i}^{2}+ y_{i}^{2}+ z_{i}^{2} \right\}}$where $x_{i},y_{i},andz_{i}$represent the acceleration samples at time index $i$. Features were extracted from the three raw axes and the calculated SVM vector. | |

Supplementary Table 2. The frequency of feature removal across the leave-one-subject-out cross-validation out of 65 subject testing. 

| Reasons for removal | Features | Frequency |
| --- | --- | --- |
| Zero Variance | Magnitude of the dominant frequency | 65 |
| High Correlation (>0.9) | Standard Deviation of the magnitude | 65 |
|  | Median of c axis | 65 |
|  | Zero crossing rate of y axis | 65 |
|  | Zero crossing rate of z axis | 65 |
|  | Range of z axis | 65 |
|  | Range of x axis | 65 |
|  | Mean of y axis | 65 |
|  | Mean of z axis | 65 |
|  | Mean magnitude | 65 |
|  | Median of magnitude | 65 |
|  | Median of y axis | 63 |
|  | Mode of z axis | 62 |
|  | Range of y axis | 62 |
|  | Mode of x axis | 43 |
|  | Mean of x axis | 22 |
|  | Median of z axis | 3 |
|  | Mode of y axis | 2 |

Supplementary Table 3. Sleep-wake performance differences for age, sex, and OSA status

| Metrics | Algorithm | term | estimate | std.error | statistic | p.value |
| --- | --- | --- | --- | --- | --- | --- |
| Balanced Accuracy | Long Short-Term Memory | Mild OSA vs No OSA | -0.01 | 0.03 | -0.44 | 0.66 |
|  |  | Moderate-Severe OSA vs No OSA | -0.01 | 0.03 | -0.37 | 0.72 |
|  |  | Age | 0.00 | 0.00 | -0.92 | 0.36 |
|  |  | Male vs Female | 0.01 | 0.02 | 0.34 | 0.73 |
|  | Random Forest | Mild OSA vs No OSA | -0.02 | 0.02 | -0.88 | 0.38 |
|  |  | Moderate-Severe OSA vs No OSA | -0.02 | 0.03 | -0.75 | 0.45 |
|  |  | Age | 0.00 | 0.00 | -1.08 | 0.28 |
|  |  | Male vs Female | 0.01 | 0.02 | 0.69 | 0.49 |
|  | Neural Network (Single layer) | Mild OSA vs No OSA | -0.01 | 0.02 | -0.55 | 0.58 |
|  |  | Moderate-Severe OSA vs No OSA | -0.04 | 0.03 | -1.40 | 0.17 |
|  |  | Age | 0.00 | 0.00 | -1.24 | 0.22 |
|  |  | Male vs Female | 0.02 | 0.02 | 1.02 | 0.31 |
|  | Logistic Regression | Mild OSA vs No OSA | 0.00 | 0.02 | -0.12 | 0.90 |
|  |  | Moderate-Severe OSA vs No OSA | -0.02 | 0.03 | -0.65 | 0.52 |
|  |  | Age | 0.00 | 0.00 | -0.66 | 0.51 |
|  |  | Male vs Female | 0.03 | 0.02 | 1.63 | 0.11 |
|  | Random Forest (Adult Model) | Mild OSA vs No OSA | -0.02 | 0.02 | -0.84 | 0.40 |
|  |  | Moderate-Severe OSA vs No OSA | 0.00 | 0.02 | -0.12 | 0.90 |
|  |  | Age | 0.00 | 0.00 | 0.73 | 0.47 |
|  |  | Male vs Female | 0.01 | 0.02 | 0.48 | 0.63 |
|  | GGIR-S | Mild OSA vs No OSA | 0.01 | 0.04 | 0.37 | 0.71 |
|  |  | Moderate-Severe OSA vs No OSA | 0.00 | 0.04 | -0.03 | 0.97 |
|  |  | Age | -0.01 | 0.01 | -1.67 | 0.10 |
|  |  | Male vs Female | -0.02 | 0.03 | -0.77 | 0.45 |
|  | GGIR-vH | Mild OSA vs No OSA | 0.02 | 0.03 | 0.51 | 0.61 |
|  |  | Moderate-Severe OSA vs No OSA | 0.02 | 0.03 | 0.56 | 0.58 |
|  |  | Age | 0.00 | 0.00 | -1.11 | 0.27 |
|  |  | Male vs Female | 0.00 | 0.03 | 0.19 | 0.85 |
|  | GGIR-CK | Mild OSA vs No OSA | 0.02 | 0.03 | 0.74 | 0.46 |
|  |  | Moderate-Severe OSA vs No OSA | 0.01 | 0.03 | 0.19 | 0.85 |
|  |  | Age | -0.01 | 0.00 | -1.52 | 0.13 |
|  |  | Male vs Female | 0.01 | 0.02 | 0.38 | 0.71 |
| Sensitivity | Long Short-Term Memory | Mild OSA vs No OSA | -0.03 | 0.03 | -1.06 | 0.29 |
|  |  | Moderate-Severe OSA vs No OSA | -0.10 | 0.04 | -2.87 | 0.01 |
|  |  | Age | 0.01 | 0.00 | 2.13 | 0.04 |
|  |  | Male vs Female | -0.02 | 0.03 | -0.64 | 0.53 |
|  | Random Forest | Mild OSA vs No OSA | -0.02 | 0.03 | -0.68 | 0.50 |
|  |  | Moderate-Severe OSA vs No OSA | -0.03 | 0.04 | -0.81 | 0.42 |
|  |  | Age | 0.01 | 0.00 | 1.31 | 0.19 |
|  |  | Male vs Female | -0.02 | 0.03 | -0.76 | 0.45 |
|  | Neural Network | Mild OSA vs No OSA | -0.02 | 0.03 | -0.59 | 0.56 |
|  |  | Moderate-Severe OSA vs No OSA | -0.07 | 0.03 | -2.02 | 0.05 |
|  |  | Age | 0.01 | 0.00 | 1.26 | 0.21 |
|  |  | Male vs Female | 0.01 | 0.02 | 0.24 | 0.81 |
|  | Logistic Regression | Mild OSA vs No OSA | 0.00 | 0.02 | -0.27 | 0.79 |
|  |  | Moderate-Severe OSA vs No OSA | -0.04 | 0.02 | -2.43 | 0.02 |
|  |  | Age | 0.01 | 0.00 | 3.18 | 0.00 |
|  |  | Male vs Female | 0.03 | 0.01 | 2.14 | 0.04 |
|  | Random Forest (Adult) | Mild OSA vs No OSA | -0.02 | 0.03 | -0.69 | 0.49 |
|  |  | Moderate-Severe OSA vs No OSA | -0.05 | 0.03 | -1.78 | 0.08 |
|  |  | Age | 0.01 | 0.00 | 1.57 | 0.12 |
|  |  | Male vs Female | 0.02 | 0.02 | 0.83 | 0.41 |
|  | GGIR-S | Mild OSA vs No OSA | 0.04 | 0.05 | 0.77 | 0.44 |
|  |  | Moderate-Severe OSA vs No OSA | 0.04 | 0.05 | 0.70 | 0.49 |
|  |  | Age | 0.00 | 0.01 | -0.07 | 0.95 |
|  |  | Male vs Female | 0.01 | 0.04 | 0.32 | 0.75 |
|  | GGIR-vH | Mild OSA vs No OSA | 0.04 | 0.04 | 0.83 | 0.41 |
|  |  | Moderate-Severe OSA vs No OSA | -0.01 | 0.05 | -0.26 | 0.79 |
|  |  | Age | 0.00 | 0.01 | 0.54 | 0.59 |
|  |  | Male vs Female | 0.03 | 0.04 | 0.80 | 0.42 |
|  | GGIR-CK | Mild OSA vs No OSA | 0.02 | 0.04 | 0.54 | 0.59 |
|  |  | Moderate-Severe OSA vs No OSA | -0.05 | 0.05 | -1.04 | 0.30 |
|  |  | Age | 0.00 | 0.01 | 0.43 | 0.67 |
|  |  | Male vs Female | 0.02 | 0.04 | 0.50 | 0.62 |
| Specificity | Long Short-Term Memory | Mild OSA vs No OSA | 0.01 | 0.05 | 0.21 | 0.84 |
|  |  | Moderate-Severe OSA vs No OSA | 0.08 | 0.06 | 1.36 | 0.18 |
|  |  | Age | -0.02 | 0.01 | -2.15 | 0.04 |
|  |  | Male vs Female | 0.03 | 0.04 | 0.70 | 0.48 |
|  | Random Forest | Mild OSA vs No OSA | -0.02 | 0.05 | -0.44 | 0.66 |
|  |  | Moderate-Severe OSA vs No OSA | -0.01 | 0.05 | -0.20 | 0.84 |
|  |  | Age | -0.01 | 0.01 | -2.15 | 0.04 |
|  |  | Male vs Female | 0.05 | 0.04 | 1.31 | 0.19 |
|  | Neural Network | Mild OSA vs No OSA | -0.01 | 0.04 | -0.19 | 0.85 |
|  |  | Moderate-Severe OSA vs No OSA | 0.00 | 0.05 | -0.10 | 0.92 |
|  |  | Age | -0.01 | 0.01 | -2.21 | 0.03 |
|  |  | Male vs Female | 0.03 | 0.04 | 0.92 | 0.36 |
|  | Logistic Regression | Mild OSA vs No OSA | 0.00 | 0.05 | -0.04 | 0.97 |
|  |  | Moderate-Severe OSA vs No OSA | 0.01 | 0.05 | 0.14 | 0.89 |
|  |  | Age | -0.01 | 0.01 | -1.75 | 0.09 |
|  |  | Male vs Female | 0.04 | 0.04 | 0.98 | 0.33 |
|  | Random Forest (Adult) | Mild OSA vs No OSA | -0.02 | 0.04 | -0.43 | 0.67 |
|  |  | Moderate-Severe OSA vs No OSA | 0.05 | 0.05 | 0.96 | 0.34 |
|  |  | Age | 0.00 | 0.01 | -0.22 | 0.82 |
|  |  | Male vs Female | 0.00 | 0.04 | -0.02 | 0.98 |
|  | GGIR-S | Mild OSA vs No OSA | -0.01 | 0.06 | -0.15 | 0.88 |
|  |  | Moderate-Severe OSA vs No OSA | -0.04 | 0.07 | -0.57 | 0.57 |
|  |  | Age | -0.02 | 0.01 | -1.92 | 0.06 |
|  |  | Male vs Female | -0.06 | 0.05 | -1.15 | 0.25 |
|  | GGIR-vH | Mild OSA vs No OSA | -0.01 | 0.05 | -0.12 | 0.90 |
|  |  | Moderate-Severe OSA vs No OSA | 0.05 | 0.06 | 0.92 | 0.36 |
|  |  | Age | -0.01 | 0.01 | -1.84 | 0.07 |
|  |  | Male vs Female | -0.02 | 0.04 | -0.48 | 0.63 |
|  | GGIR-CK | Mild OSA vs No OSA | 0.01 | 0.04 | 0.36 | 0.72 |
|  |  | Moderate-Severe OSA vs No OSA | 0.06 | 0.04 | 1.44 | 0.15 |
|  |  | Age | -0.01 | 0.01 | -2.50 | 0.02 |
|  |  | Male vs Female | 0.00 | 0.03 | -0.07 | 0.94 |

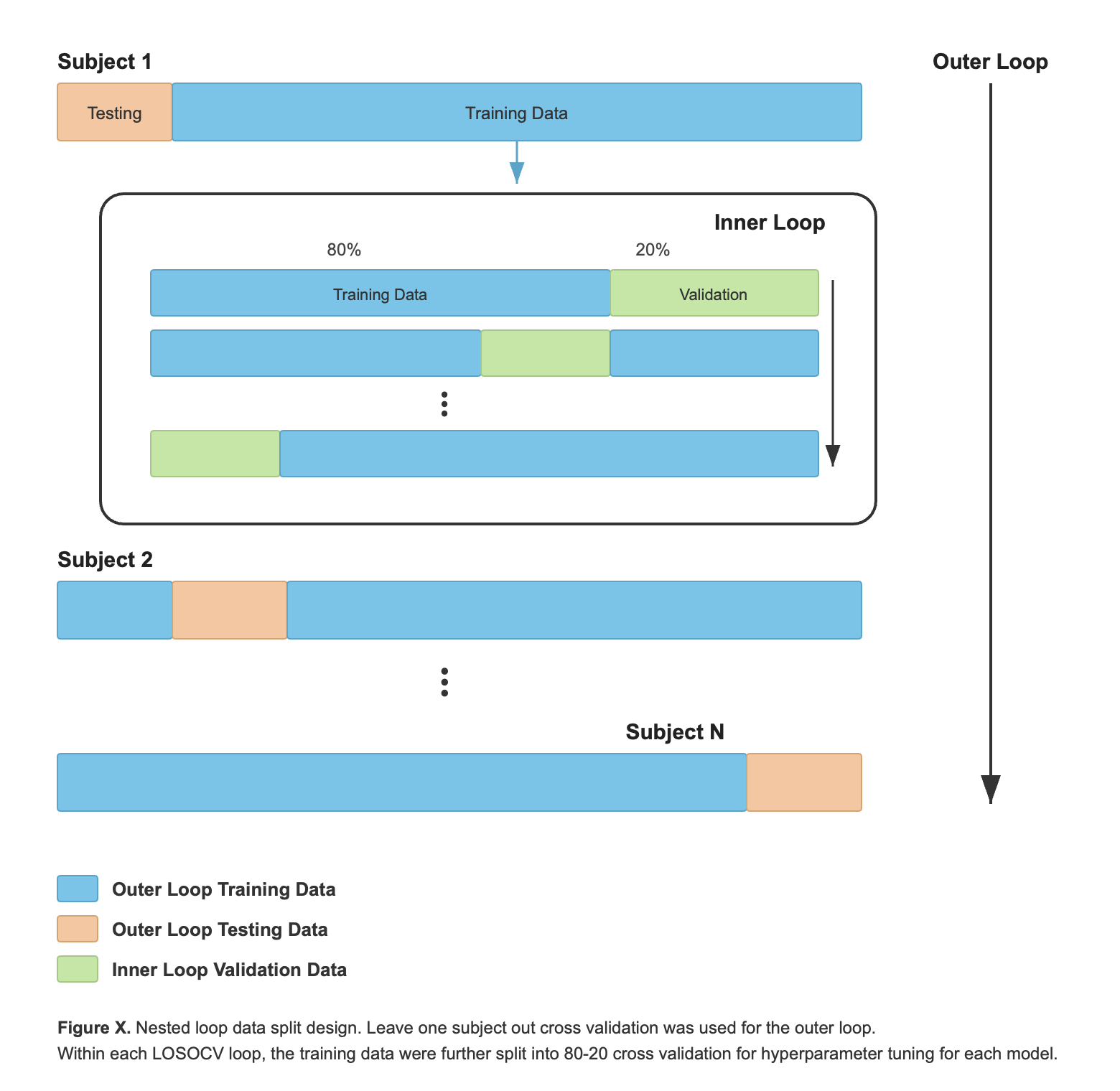

Supplementary Figure 1. Nested loop data split design. Leave-one-subject-out cross-validation was used for the outer loop. Within each LOSOCV loop, the training data were further split into an 80-20 cross-validation scheme for hyperparameter tuning for each model.

Supplementary Figure 2. Random Forest Sleep duration Bland-Altman plot

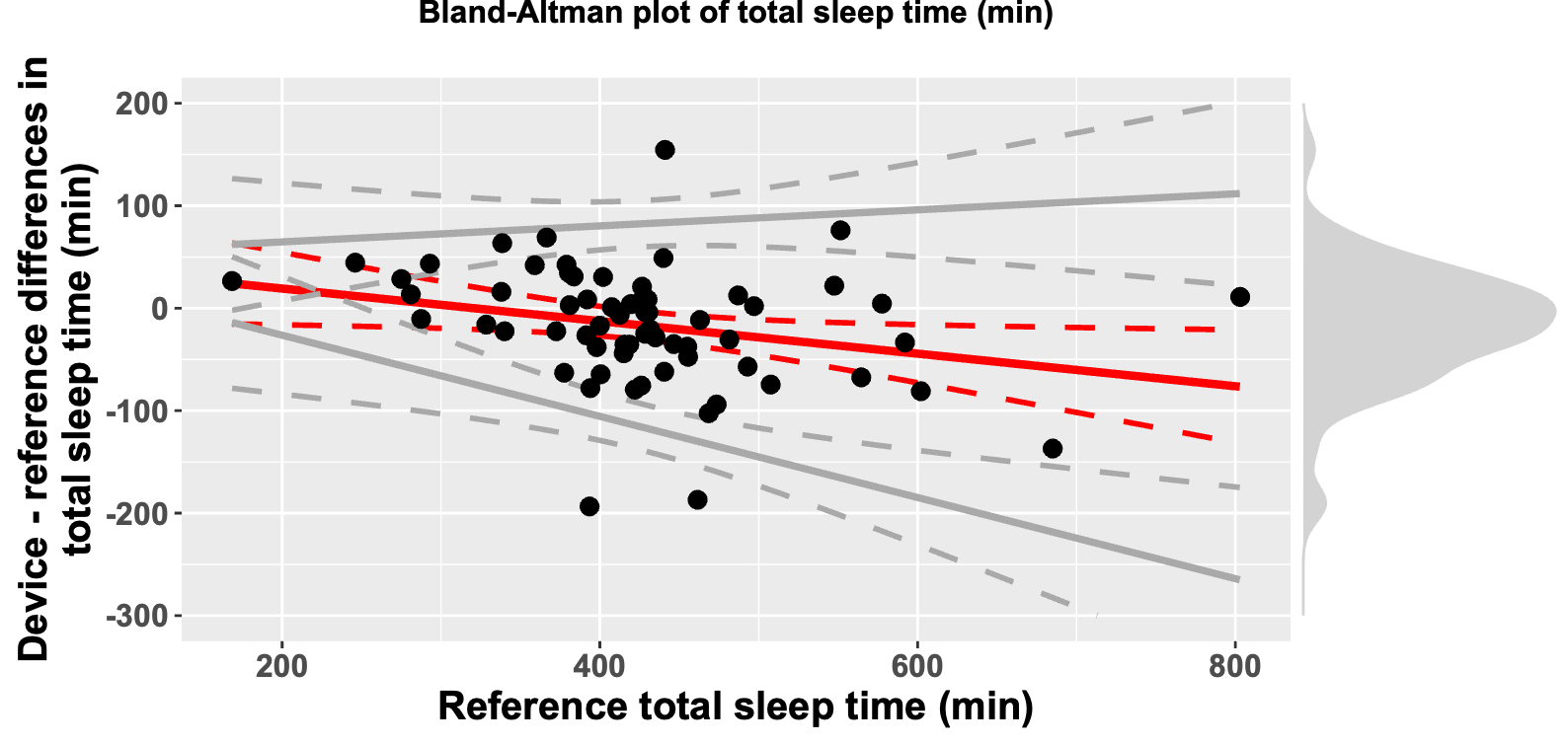

Supplementary Figure 3. Random Forest WASO Bland-Altman plot

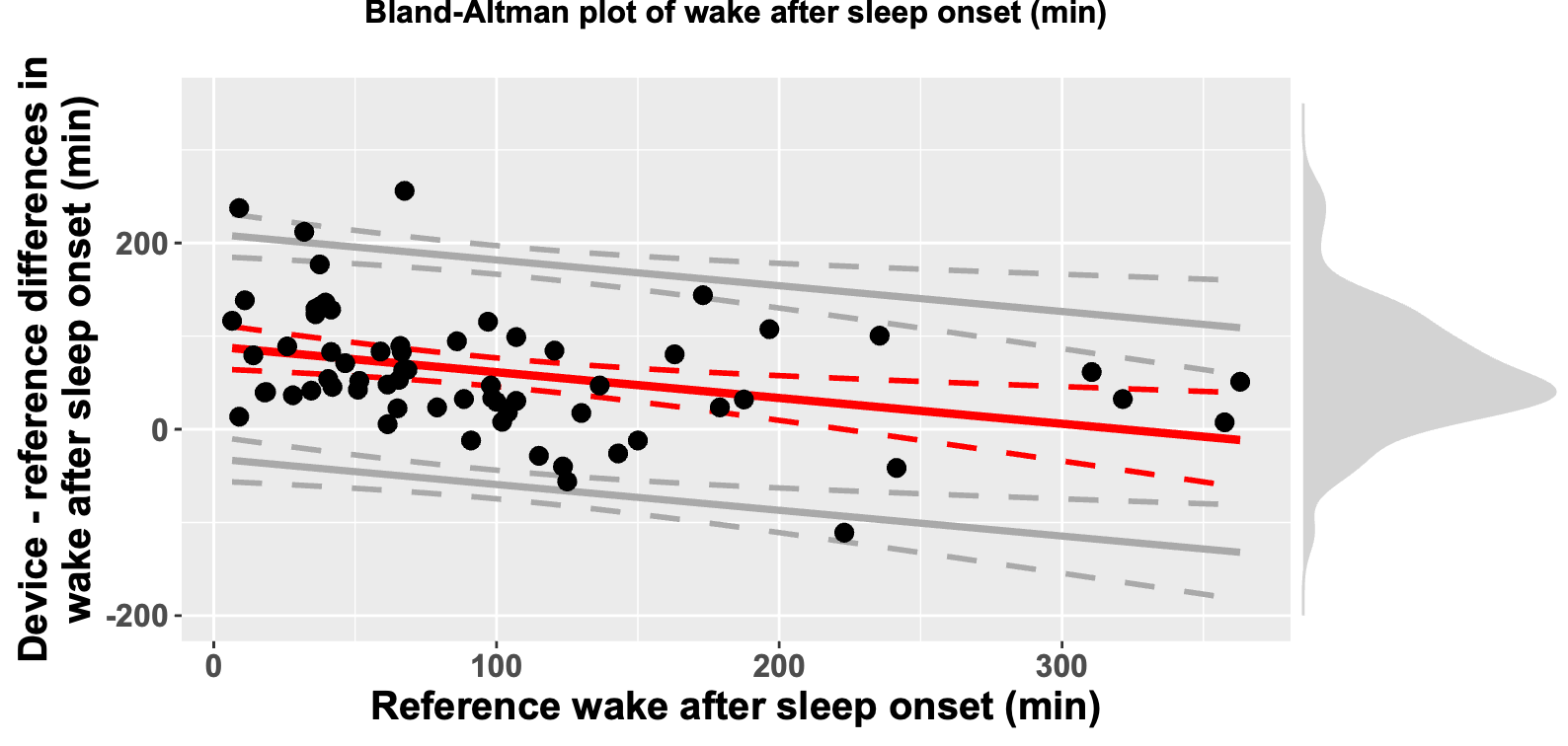

Supplementary Figure 4. Neural Network Sleep Duration Bland-Altman plot

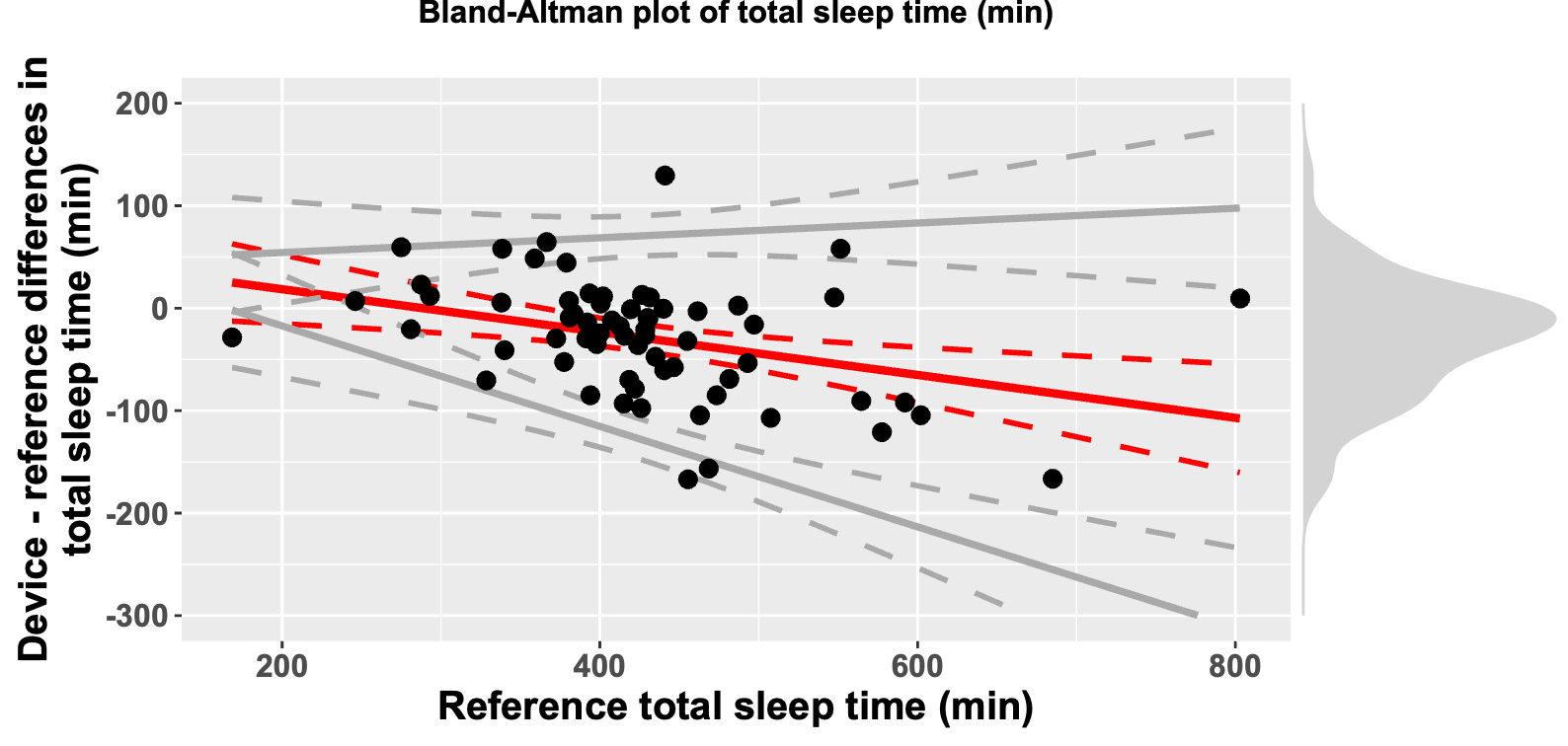

Supplementary Figure 5. Neural Network WASO Bland-Altman plot

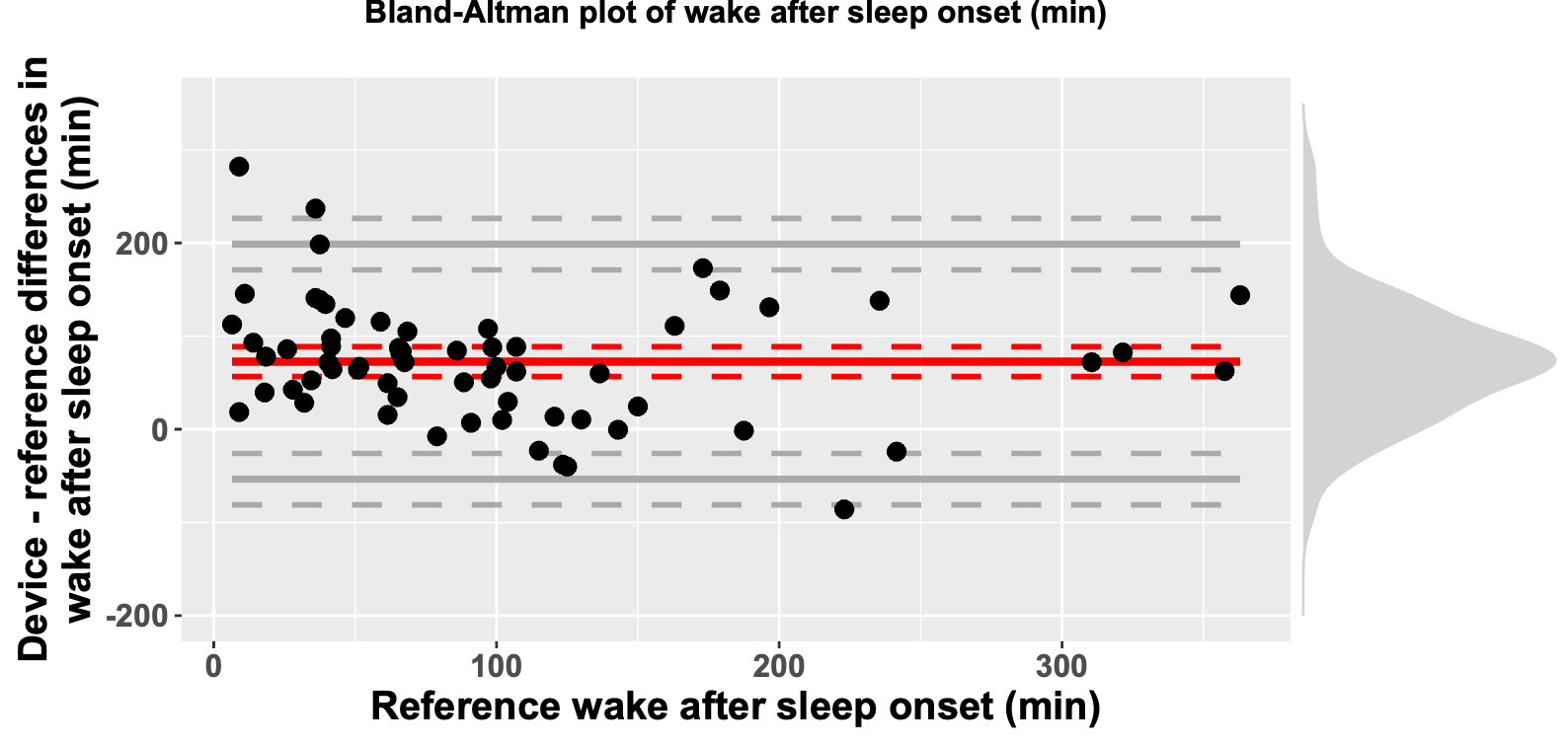

Supplementary Figure 6. Logistic Regression Sleep Duration Bland-Altman plot

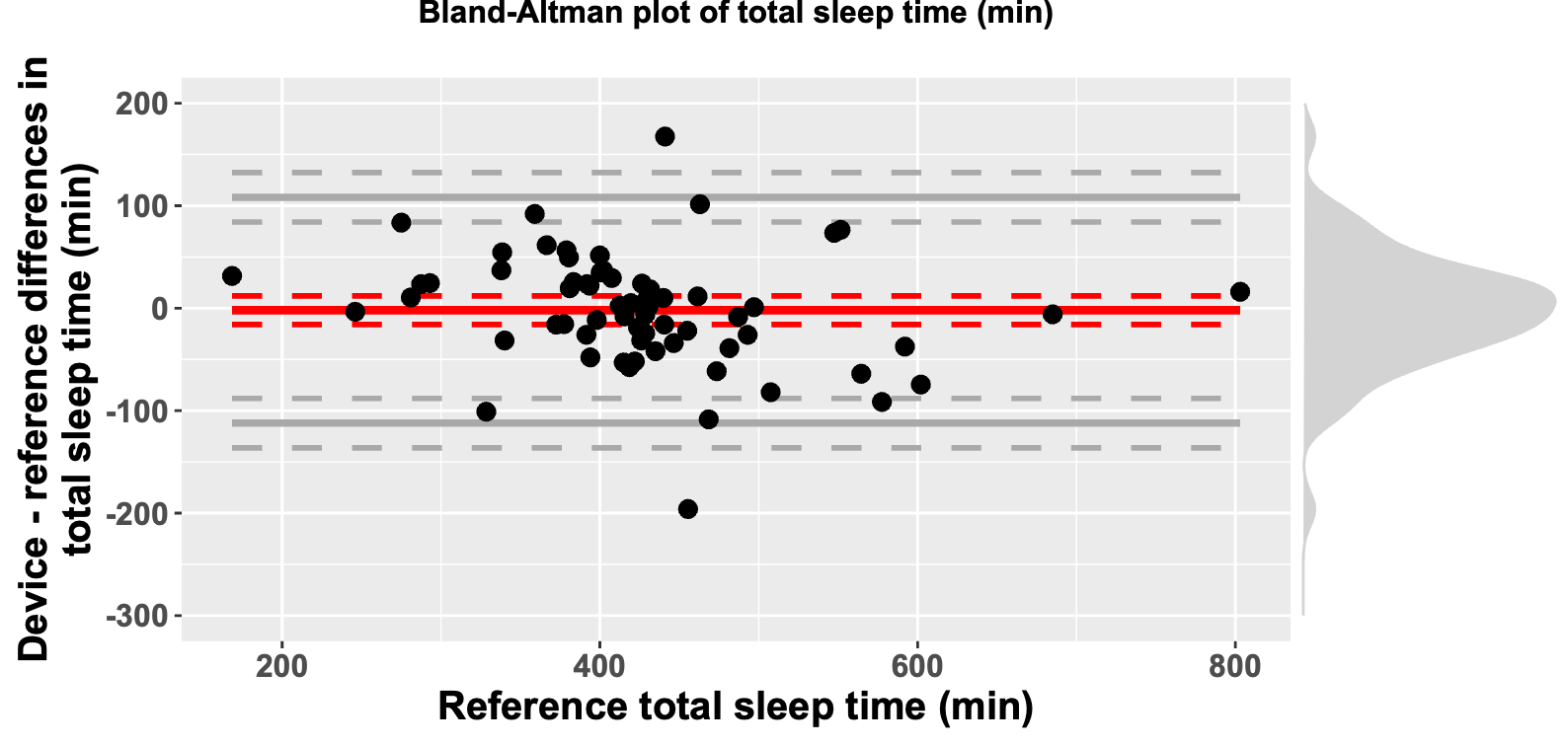

Supplementary Figure 7. Logistic Regression WASO Bland-Altman plot

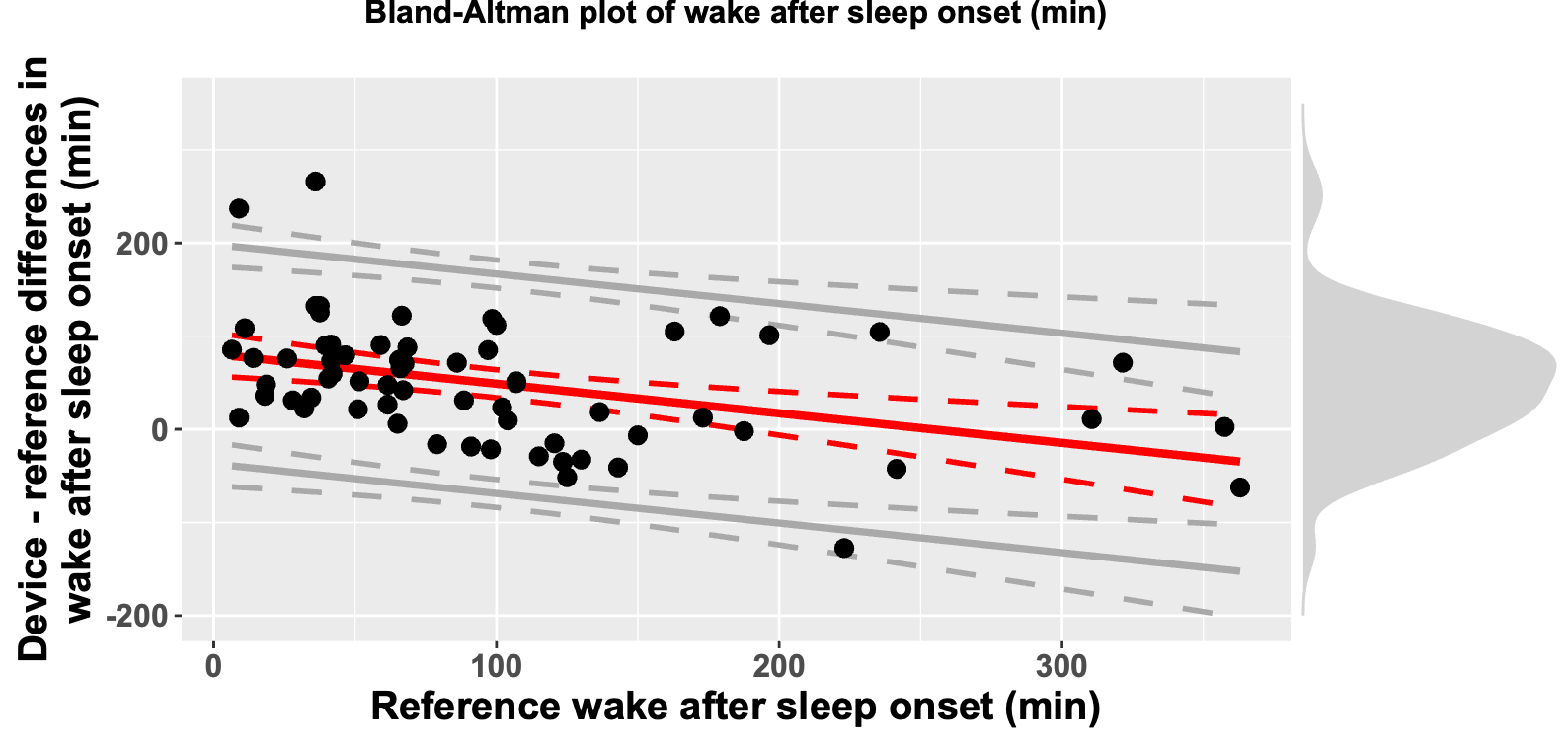

Supplementary Figure 8. Long Short Term Memory algorithm Sleep Duration Bland-Altman plot

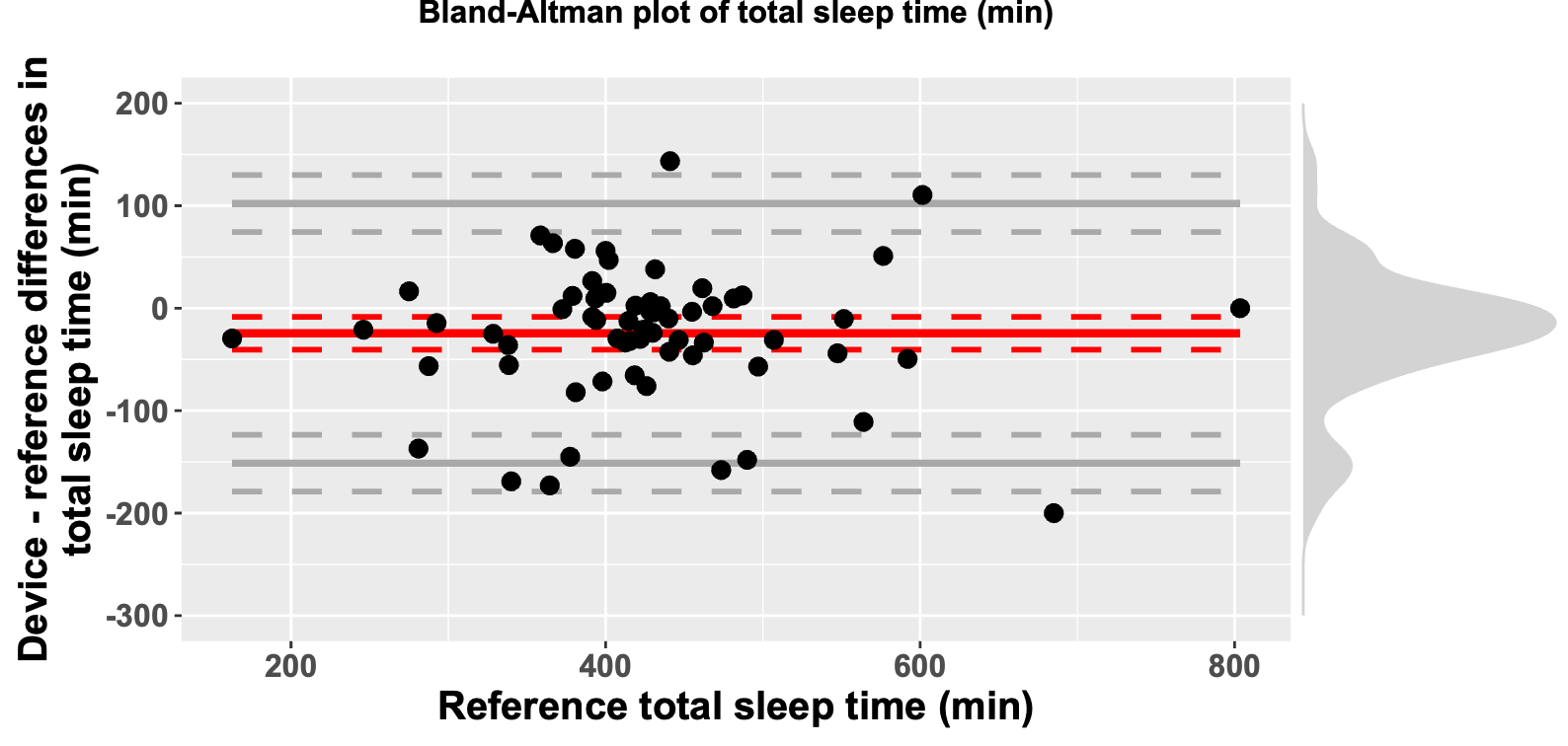

Supplementary Figure 9. Long Short Term Memory algorithm WASO Bland-Altman plot

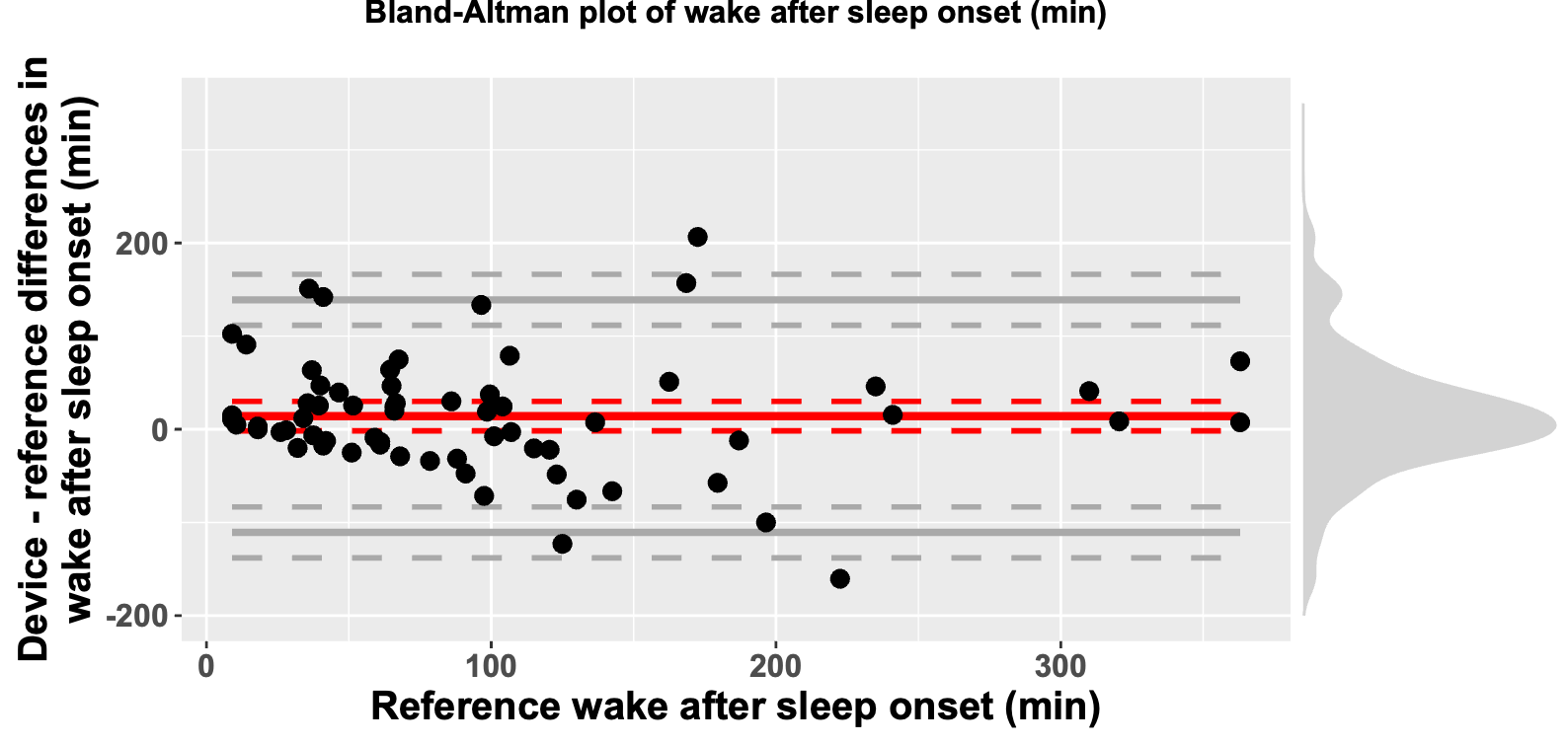

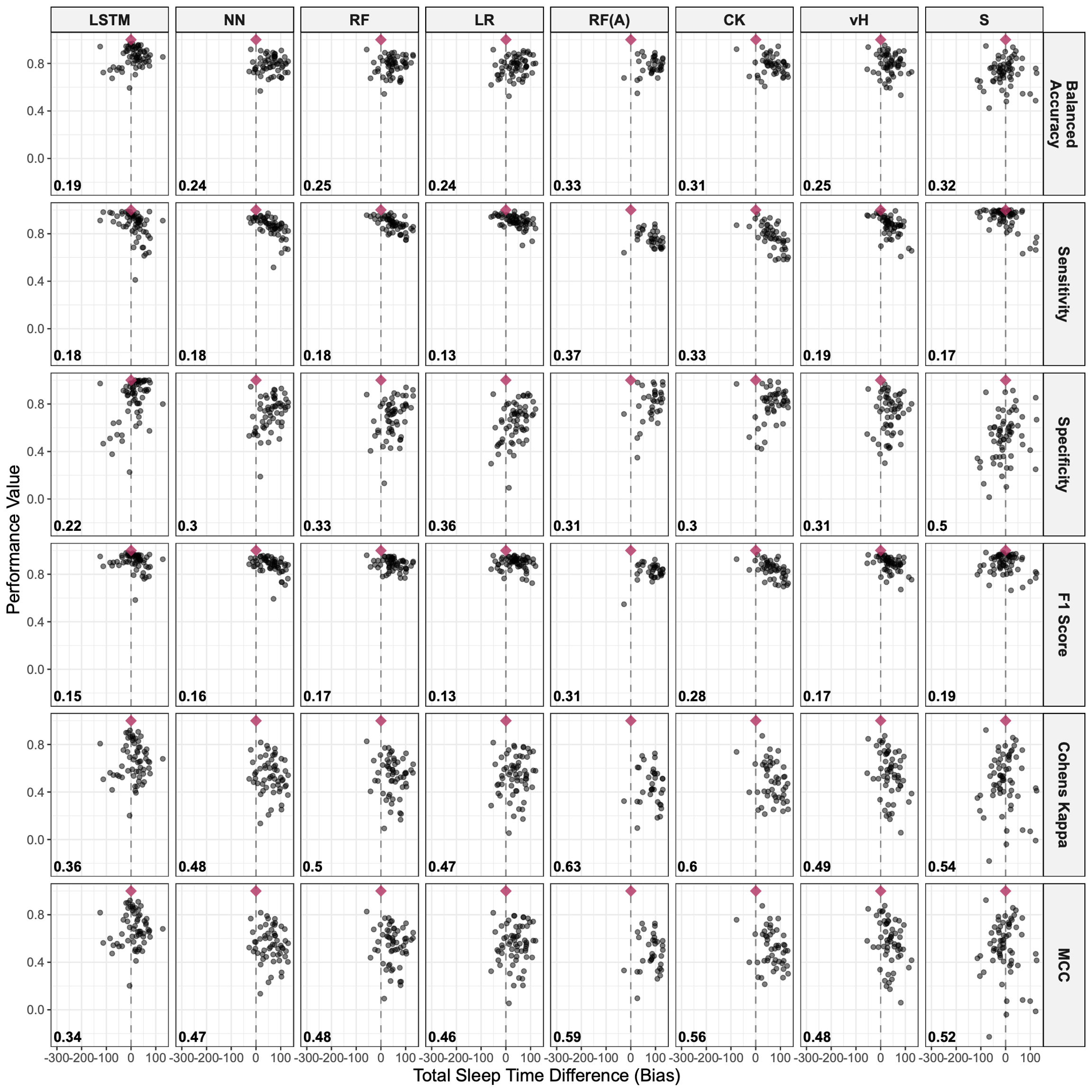

Supplementary Figure 10. Scatter plot of WASO bias versus performance metrics. The red diamond indicates a perfect score (0 minutes of bias and a metric score of 1). The dashed line represents 0 minutes of bias. Tighter clustering indicates higher model reliability, while proximity to the red diamond signifies better overall model performance. LSTM = Long Short-Term Memory, NN = Neural Network, RF = Random Forest, LR = Logistic Regression, RF(A) = Random Forest (Adult), CK = GGIR-adapted Cole-Kripke, vH = GGIR-adapted van Hees, S = GGIR- adapted Sadeh.

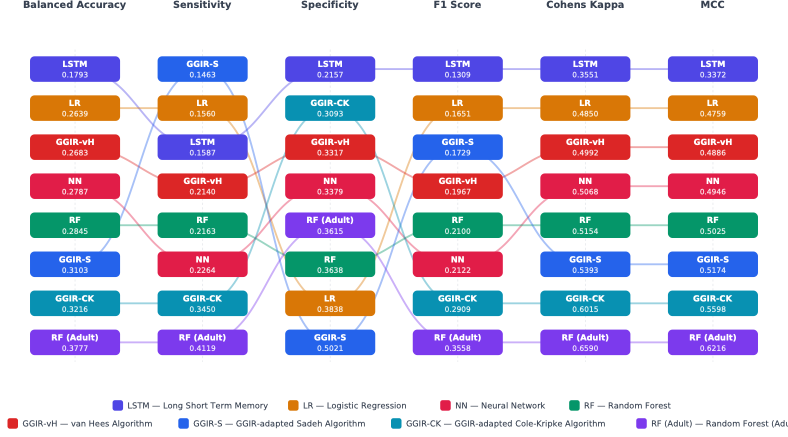

Supplementary Figure 11. Sankey diagram of Euclidean Composite score for the WASO bias and balanced accuracy score of the top machine learning and benchmark references.
